# Vaccine effectiveness of the first dose of ChAdOx1 nCoV-19 and BNT162b2 against SARS-CoV-2 infection in residents of Long-Term Care Facilities (VIVALDI study)

**DOI:** 10.1101/2021.03.26.21254391

**Authors:** Madhumita Shrotri, Maria Krutikov, Tom Palmer, Rebecca Giddings, Borscha Azmi, Sathyavani Subbarao, Christopher Fuller, Aidan Irwin-Singer, Daniel Davies, Gokhan Tut, Jamie Lopez Bernal, Paul Moss, Andrew Hayward, Andrew Copas, Laura Shallcross

## Abstract

**Background:** The effectiveness of SARS-CoV-2 vaccines in frail older adults living in Long-Term Care Facilities (LTCFs) is uncertain. We estimated protective effects of the first dose of ChAdOx1 and BNT162b2 vaccines against infection in this population.

**Methods:** Cohort study comparing vaccinated and unvaccinated LTCF residents in England, undergoing routine asymptomatic testing (8 December 2020 - 15 March 2021). We estimated the relative hazard of PCR-positive infection using Cox proportional hazards regression, adjusting for age, sex, prior infection, local SARS-CoV-2 incidence, LTCF bed capacity, and clustering by LTCF.

**Results:** Of 10,412 residents (median age 86 years) from 310 LTCFs, 9,160 were vaccinated with either ChAdOx1 (6,138; 67%) or BNT162b2 (3,022; 33%) vaccines. A total of 670,628 person days and 1,335 PCR-positive infections were included. Adjusted hazard ratios (aHRs) for PCR-positive infection relative to unvaccinated residents declined from 28 days following the first vaccine dose to 0·44 (0·24, 0·81) at 28-34 days and 0·38 (0·19, 0·77) at 35-48 days. Similar effect sizes were seen for ChAdOx1 (aHR 0·32 [0·15-0·66] and BNT162b2 (aHR 0·35 [0·17, 0·71]) vaccines at 35-48 days. Mean PCR cycle threshold values were higher, implying lower infectivity, for infections ≥28 days post-vaccination compared with those prior to vaccination (31·3 vs 26·6, p<0·001).

**Interpretation:** The first dose of BNT162b2 and ChAdOx1 vaccines was associated with substantially reduced SARS-CoV-2 infection risk in LTCF residents from 4 weeks to at least 7 weeks.

**Funding:** UK Government Department of Health and Social Care.

**Research in Context:** *Evidence before this study:* We conducted a systematic search for studies which evaluated SARS-CoV-2 vaccine effectiveness in residents of long-term care facilities (LTCFs) published between 01/01/2020 and 11/03/2021. We used variations of search terms for “COVID-19” AND “vaccine effectiveness” OR “vaccine efficacy” AND “care homes” OR “long term care facilities” OR “older people” on Ovid MEDLINE and MedRxiv. We identified one pre-print article regarding LTCFs in Denmark, which reported that a single dose of BNT162b was ineffective against SARS-CoV-2 infection in residents, however, participants received the second vaccine dose 24 days following the first dose on average, which is likely to be too soon to capture the protective effects of a single vaccine dose. Additionally, we identified two pre-print reports of studies evaluating vaccine effectiveness against symptomatic infection and hospitalisation amongst older adults in the community. The first of these found 81% vaccine effectiveness against COVID-19-related hospitalisation at 28-34 days following a single dose of BNT162b or ChAdOx1 in ≥80-year-olds. The second of these found vaccine effectiveness against symptomatic infection of 60% at 28-34 days and 73% at 35+ days following a single dose of ChAdOx1 in ≥70-year-olds. No studies were identified that focused on the effectiveness of a single vaccine dose against infection amongst LTCF residents at more than 4 weeks post-vaccination, a particularly important question in the context of the UK policy decision to extend the dose interval beyond 3 weeks.

*Added value of this study:* We conducted a prospective cohort study of 10,412 residents aged ≥65 years, from 310 LTCFs across England, to investigate the protective effect of the first dose of the ChAdOx1 and BNT162b vaccines against SARS-CoV-2 infection in frail older adults. We retrieved results from routine monthly PCR testing, as well as outbreak and clinical testing for SARS-CoV-2, thereby capturing data on asymptomatic as well as symptomatic infections, which we linked to vaccination records. We estimated vaccine effectiveness to be 56% (19-76%) at 28-34 days, and 62% (23-81%) at 35-48 days following a single dose of ChAdOx1 or BNT162. Our findings suggest that the risk of SARS-CoV-2 infection is substantially reduced from 28 days following the first dose of either vaccine and that this effect is maintained for at least 7 weeks, with similar protection offered by both vaccine types. We also found that PCR cycle threshold (Ct) values, which are negatively associated with the ability to isolate virus, were significantly higher in infections occurring at ≥ 28days post vaccination compared to those occurring in the unvaccinated period, suggesting that vaccination may reduce onward transmission of SARS-CoV-2 in breakthrough infections. To the best of our knowledge, our findings constitute the first real-world evidence on vaccine effectiveness against infection for ChAdOx1, in any age group. We can also infer that both vaccines are effective against the B.1.1.7 variant, because our analysis period coincided with the rapid emergence of B.1.1.7 in England during the second wave of the pandemic.

*Implications of all the available evidence:* Our findings add to the growing body of evidence on the protective effect of the BNT162b vaccines in residents of LTCFs and demonstrate the effectiveness of ChAdOx1 in this vulnerable population. Evaluating single-dose vaccine efficacy has become increasingly important in light of extended dosing intervals that have been implemented in order to maximise vaccine coverage across high-risk groups. Further work is required to evaluate the effectiveness of the first vaccine dose after 8-12 weeks, as well as following the second dose, and to evaluate the long-term impact of vaccination on SARS-CoV-2 infection, transmission and mortality in LTCFs. This will inform policy decisions regarding the ongoing need for disease control measures in LTCF such as visitor restrictions, which continue to have a detrimental impact on the wellbeing of residents, their relatives, and staff. Supplementary material attached.

## Introduction

The greatest impact of Severe Acute Respiratory Syndrome Coronavirus 2 (SARS-CoV-2) has been seen in residents of Long-Term Care Facilities (LTCFs), who represent less than 1% of the population, ^1^ but account for a large proportion of SARS-CoV-2 related deaths in European and North American countries.^2–4^ The UK has prioritised vaccination of residents and staff in LTCFs^5^ to reduce the risk of Coronavirus disease 2019 (COVID-19)-related morbidity and mortality in this population, with the expectation that this will facilitate the relaxation of social restrictions.

Two vaccines have been deployed in LTCFs in England: Oxford/AstraZeneca’s non-replicating viral-vectored vaccine (ChAdOx1 nCoV-19), licensed on 30 December 2020,^6^ and Pfizer/BioNTech’s messenger RNA-based vaccine (BNT162b2), licensed on 2 December 2020.^7^ Both are Spike-protein based vaccines showing high efficacy (62·1-95·0%) in Phase 3 clinical trials when following a two-dose schedule.^8,9^ However, trials for both vaccines have enrolled mostly younger, healthy adults. Vaccine efficacy data from frail older adults requiring long-term care are limited because these individuals are routinely excluded from clinical studies and vaccine trials.^10,11^ Consequently, trial estimates of vaccine efficacy may not be generalisable to LTCF residents due to age-related differences in vaccine-induced immune responses.^12–14^ Observational data from post-licensure studies in older adults are emerging,^15,16^ but few studies report specifically on LTCF populations.

Manufacturer-recommended intervals between first and second doses of BNT162b and ChAdOx1 vaccines are three and four weeks, respectively, though trial data indicates higher efficacy with an extended dosing interval for ChAdOx1.^17^ On 31 December 2020 the UK Joint Committee on Vaccination and Immunisation advised that the intervals could be extended up to 12 weeks,^18^ in order to optimise first dose coverage. This decision was taken in the context of rapidly increasing SARS-CoV-2 incidence, associated with the emergence of the highly transmissible B.1.1.7 variant and its subsequent spread within LTCFs from November 2020.^19,20^ This policy has made it increasingly important to understand the extent and duration of protection against infection afforded by the first dose of each vaccine, and whether single dose vaccination impacts on transmission. This knowledge will inform re-vaccination schedules as well as policy decisions regarding the ongoing need for LTCF specific control measures which aim to prevent transmission, such as visitor restrictions and asymptomatic testing.

As all LTCF residents aged ≥ 65 years have now been offered one dose of the vaccine, we analysed data from our prospective observational cohort study to investigate the protective effect of the first dose of ChAdOx1 and BNT162b2 vaccines in this population. We compared the relative hazards of PCR-confirmed SARS-CoV-2 infections and mean PCR Cycle Threshold (Ct) values in vaccinated and unvaccinated residents by time since vaccination.

## Methods

The VIVALDI study is a prospective cohort study, which was set up in May 2020 to investigate SARS-CoV-2 transmission, infection outcomes, and immunity in residents and staff in LTCFs in England that provide residential and/or nursing care for adults aged 65 years and over.^21^ ‘For-profit’ and ‘not-for-profit’ chain Providers, and independent Providers of differing sizes from all regions across England are participating in the study. Eligible LTCFs were identified by the Care Provider’s Senior Management Team, or by the National Institute for Health Research (NIHR) Clinical Research Network. Pseudonymised clinical and demographic data were retrieved for staff and residents from participating LTCFs through national surveillance systems.

Since June 2020 all residents in LTCFs in England are offered regular SARS-CoV-2 testing using polymerase chain reaction (PCR)-based assays of nasopharyngeal swab specimens.^22^ LTCF residents undergo monthly routine PCR testing, and if an LTCF outbreak is suspected, local public health teams organise PCR testing for all residents upon notification and 7 days later. Individuals who test positive are not re-tested for the following 90 days unless they develop new COVID-19 symptoms.^23^ Symptom information is collected at the point of testing but its reliability is uncertain in this frail population. Routine PCR testing data, including both positive and negative results, as well as any positive PCR results from hospital-based clinical testing, were retrieved from the COVID-19 Datastore [https://data.england.nhs.uk/covid-19/], which was established as part of the UK’s pandemic response. Void tests were excluded from the analysis.

Age, sex, and LTCF location were obtained for all participants from national surveillance datasets. Vaccination records (date, vaccine type, dose number) were retrieved from the National Immunisation Management Service (NIMS). Estimates of the total number of beds and weekly bed occupancy in each LTCF were retrieved from Capacity Tracker [https://www.necsu.nhs.uk/capacity-tracker], a national reporting tool collating weekly operational data from LTCF managers. England is divided into 343 local authorities which are responsible for the delivery of community services such as social care and education. Weekly SARS-CoV-2 incidence estimates at local authority level, produced by the Department of Health and Social Care [https://www.gov.uk/government/collections/coronavirus-cases-by-local-authority-epidemiological-data], were used to derive the average monthly incidence of SARS-CoV-2 in the area surrounding each LTCF. Subject to informed consent, blood sampling to detect IgG antibodies to Nucleocapsid protein was offered to a subset of residents prior to vaccination. Where available, Cycle threshold (Ct) values were retrieved for positive PCR tests within the study period.

### Data linkage

PCR results from the national testing programme can be linked to specific residents using a pseudo-identifier which is based on the individuals’ unique National Health Service (NHS) number. The Care Quality Commission regulates all providers of health and social care in England. PCR results from the national testing programme are linked to specific care homes using the Care Quality Commission’s unique location ID (CQC-ID), making it possible to link residents to specific LTCFs.

We retrieved PCR test results, and associated data on age, sex, and CQC-ID, from 1 March 2020 onwards. Residents with PCR tests linked to multiple CQC-IDs were assigned to the LTCF corresponding to their most recent PCR test. The NHS number-based pseudo-identifier was used to retrieve vaccination records from the NIMS database, and to link to antibody test results, which are both held in the COVID-19 Datastore (Supplement 1). Vaccination data from NIMS were validated against coverage estimates from LTCFs where available. LTCFs with no record of resident vaccination within NIMS were excluded from the analysis. The linked dataset was analysed in the UCL Data Safe Haven.

### Population

We conducted an individual level analysis of the risk of PCR-confirmed SARS-CoV-2 infection by vaccination status amongst residents aged 65 years and over, from LTCFs enrolled in the VIVALDI study. The analysis period started from the date of first vaccination in the cohort (8 December 2020) and ended at the date of last data extraction (15 March 2021). Residents were eligible for inclusion if they had at least two PCR test results in total, and ≥ 1 PCR result during the analysis period. Residents entered the risk period on 8 December 2020 if they had ≥ 1 valid PCR result on or prior to that date; or, if they had no PCR results before 8 December 2020, on the date of their first negative PCR test. Residents with a positive PCR result ≤ 90 days before 8 December entered the risk period 90 days after their positive test. Residents exited the risk period at the earliest of the following events: positive PCR test; date of second vaccination, last available PCR test.

We conducted a sensitivity analysis removing individuals who were never vaccinated despite having a PCR test more than 30 days following the date of first vaccination in their LTCF to account for potential bias from systematic differences in clinical or other features of this group.

### Outcome

The primary outcome was time to first positive PCR test, indicating SARS-CoV-2 infection, within the analysis period.

### Exposures

Vaccination status was included as a time-varying covariate using the following exposure categories: unvaccinated, and 0-6 days, 7-13 days, 14-20 days, 21-27 days, 28-34 days, 35-48 days, and 49 or more days following the first dose of either vaccine.

### Covariates

Positive PCR results from before the analysis period, and positive anti-Nucleocapsid antibody results prior to vaccination, were combined into a binary variable for prior SARS-CoV-2 infection. The average monthly incidence of SARS-CoV-2 infection in the surrounding area to each LTCF was estimated by averaging weekly incidence rates in the corresponding local authority. Bed capacity was defined as the total number of beds in each LTCF, as reported in Capacity Tracker or directly by LTCFs. The mean cycle threshold (Ct) value for each PCR-positive test was calculated by taking the mean of the available Ct values from up to 3 gene targets (Nucleocapsid, ORF1ab, Spike) for each sample (Table S1).

### Statistical Analysis

We used Cox proportional hazards models to derive hazard ratios for the risk of first PCR-positive test in the study period, comparing post-vaccination exposure categories to the unvaccinated group, with 95% confidence intervals (CI) calculated using robust standard errors to account for the non-independence of infection events within LTCFs. The baseline hazard was defined over calendar time. We adjusted for sex (as a binary variable), age (as a cubic spline term), evidence of prior SARS-CoV-2 infection (as a binary variable), LTCF bed capacity (as a linear term), and average monthly SARS-CoV-2 incidence rate for the local authority in which the LTCF was located (as a linear term). In secondary analyses we explored vaccine effects stratified by type of vaccine (ChAdOx1 and BNT162b2) and by evidence of prior SARS-CoV-2 infection. Our stratified analyses were conducted by including an interaction term between the time varying exposure status and the stratifying factor. We calculated vaccine effectiveness as 1 – the adjusted hazard ratio for infection. We used two-tailed t-tests to estimate the difference in mean Ct values between unvaccinated and vaccinated groups.

All analyses were pre-specified in a statistical analysis plan and were conducted in Stata version 16·0 (StataCorp, 2019, TX, USA).

### Sample Size and Power Considerations

Sample size for the VIVALDI study was based on the precision of estimates for antibody prevalence,^21^ therefore *a priori* sample size calculations were not performed for this analysis.

### Ethical Approval

Ethical approval for the study was obtained from the South Central - Hampshire B Research Ethics Committee, Ref: 20/SC/0238.

## Results

A total of 10,412 care home residents aged ≥65 years from 310 LTCFs were included in this analysis (Table 1). The median age was 86 years (IQR 80-91), 7,247 (69·6%) were female, and 1,155 residents (11·1%) had evidence of prior SARS-CoV-2 infection (194 seropositive, 1,013 PCR-positive). Overall, 9,160 (88·0%) residents had received at least one vaccine dose, of whom 6,138 (67·0%) received ChAdOx1 and 3,022 (33·0%) received BNT162b2. 897 participants had also received a second vaccine dose and the median dose interval was 63 days (IQR 55-65).

**Table 1.** Characteristics of individual LTCF residents included in the analysis.

| **LTCF Residents** | *n* | % |
| --- | --- | --- |
| Total | 10,412 | NA |
| Age (years) | 86 | IQR 80-91 |
| Female sex | 7,247 | 69·6% |
| Linked with single LTCF | 10,271 | 98·6% |
| **Evidence of prior infection** |  |  |
| Pre-vaccination N-antibody result available | 864 | 8·3% |
| N-antibody positive pre-vaccination | 194 | 22·5% |
| PCR-positive prior to analysis period | 1,013 | 9·7% |
| Total with evidence of prior infection | 1,155 | 11·1% |
| **PCR Tests** |  |  |
| Total PCR results in at risk period | 36,352 | NA |
| - Routine PCR tests | 36,144 | 99·4% |
| - Symptomatic at time of routine testing | 246 | 0·6% |
| PCR results per person per month | Median: 1·6 | IQR: 1·2-2·2 |
| PCR-positive events in analysis period | 1,335 | 3·7% |
| - Routine PCR tests | 1,128 | 84·5% |
| - Symptomatic at time of routine testing | 84 | 7·5% |
| **Vaccination** |  |  |
| First vaccine dose | 9,160 | 88·0% |
| ChAdOx1 | 6,138 | 67·0% |
| BNT162b2 | 3,022 | 33·0% |
| Second vaccine dose | 897 | 8·6% |
| ChAdOx1 | 179 | 20·0% |
| BNT162b2 | 718 | 80·0% |
| Different first and second dose vaccine types | 3 | 0·3% |
| Dose interval (days) | Median: 63 | IQR: 55-65 |

NA – not applicable

Data were available from 228 for-profit and 72 not-for-profit chain Providers, and 10 independent Facilities (Table 2), with a median LTCF capacity of 48 beds (IQR 40-63). Most (>75%) included LTCFs had commenced resident vaccinations by 16 January 2021 and completed most of these within two days. LTCFs which mainly used ChAdOx1 (>75% vaccinations) tended to be slightly smaller than Facilities that mainly used BNT162b (median bed capacity 47, IQR: 38-61 versus 51, IQR: 42-64); they also started vaccinations slightly later (19 versus 7 January 2021).

**Table 2.** Characteristics of LTCFs included in the analysis

| **Long-Term Care Facilities** | *n* | % |
| --- | --- | --- |
| Total LTCFs | 310 | NA |
| For-profit chain | 228 | 73.6% |
| -No. of included residents | 7,456 | 71.6% |
| Not-for-profit chain | 72 | 23.2% |
| -No. of included residents | 2,537 | 24.4% |
| Independent | 10 | 3.2% |
| -No. of included residents | 419 | 4.0% |
| Total bed capacity | Median: 48 | IQR: 40-63 |
| Occupied beds | Median: 38.5 | IQR: 30.0-50.2 |
| Occupancy | Median: 82.5% | IQR: 71.7-90.5% |
| Vaccinated residents | Median: 90.9% | IQR: 85.7-95.7% |
| **Mostly (>75%) ChAdOx1 resident vaccinations** | 203 | 65.5% |
| Total bed capacity | Median: 47 | IQR: 38-61 |
| Occupied beds | Median: 37 | IQR: 28-48 |
| Date by which >75% LTCFs started vaccination | 19 Jan 2021 | NA |
| Date by which >75% LTCFs completed >75% vaccinations | 21 Jan 2021 | NA |
| Days taken to complete >75% vaccinations | Median: 1 | IQR: 1-2 |
| **Mostly (>75%) BNT162b2 resident vaccinations** | 99 | 31.9% |
| Total bed capacity | Median: 51 | IQR: 42-64 |
| Occupied beds | Median: 41 | IQR: 32-54 |
| Date by which >75% LTCFs started vaccination | 7 Jan 2021 | NA |
| Date by which >75% LTCFs completed >75% vaccinations | 8 Jan 2021 | NA |
| Days taken to complete >75% vaccinations | Median: 1 | IQR: 1-1 |

NA – not applicable

Between 8 December 2020 and 9 March 2021, there were a total of 36,352 PCR results in 670,628 person days (54·2 PCR tests per 1,000 person days) (Table 3). The vast majority of PCR results (36,144, 99·4%) were completed as part of routine testing and the median number of PCR tests per resident per month was 1·6 (IQR 1·2-2·2). Of 1,335 positive PCR tests, 1,128 were from routine testing (84·5%) (Table 1). The crude infection rate per 10,000 person days at risk was 19·9 overall and 21·4 within the unvaccinated group (Table 3). In vaccinated residents the infection rate reduced to 9·7 and 9·4 per 10,000 person days at 28-34 and at 35-48 days post-vaccination, respectively.

**Table 3.** Infection rates and adjusted hazard ratios* for PCR-positive infection compared with unvaccinated group, for the first dose of either vaccine, by days since vaccination.

|  | Person days at risk | PCR tests | PCR testing rate per 1,000 person days | Infection events | Infection rate per 10,000 person days | Adjusted Hazard Ratio | aHR 95% CI |  | p-value |
| --- | --- | --- | --- | --- | --- | --- | --- | --- | --- |
| Unvaccinated | 338,003 | 15,392 | 45·54 | 723 | 21·39 | 1 | NA | NA | NA |
| 0-6 days | 47,591 | 2,482 | 52·15 | 105 | 22·06 | 0·64 | 0·38 | 1·06 | 0·083 |
| 7-13 days | 53,511 | 3,189 | 59·60 | 139 | 25·98 | 0·83 | 0·54 | 1·28 | 0·404 |
| 14-20 days | 50,362 | 2,462 | 48·89 | 132 | 26·21 | 0·96 | 0·57 | 1·60 | 0·866 |
| 21-27 days | 47,514 | 2,478 | 52·15 | 95 | 19·99 | 0·92 | 0·53 | 1·59 | 0·762 |
| 28-34 days | 43,136 | 2,078 | 48·17 | 42 | 9·74 | 0·44 | 0·24 | 0·81 | 0·009 |
| 35-48 days | 63,012 | 4,681 | 74·29 | 59 | 9·36 | 0·38 | 0·19 | 0·77 | 0·007 |
| 49+ days | 27,499 | 3,590 | 130·55 | 40 | 14·55 | 0·49 | 0·20 | 1·17 | 0·108 |
| Overall | 670,628 | 36,352 | 54·21 | 1,335 | 19·91 | NA | NA | NA | NA |

NA – not applicable
\*Adjusted hazard ratios were estimated using a Cox proportional hazards regression model with an interaction term between vaccination status and prior infection status; the comparator is the unvaccinated group with evidence of prior infection. Hazard ratios are adjusted for age, sex, local monthly infection incidence, LTCF bed capacity, and for estimates from the group with evidence of prior infection. 95% confidence intervals were calculated using robust standard errors for LTCF-level effects.

In Cox regression comparing vaccinated and unvaccinated groups and adjusting for age, sex, prior infection, LTCF bed capacity, and local incidence of infection, there was no significant reduction in the adjusted hazard ratios for PCR-positive infection until 28-34 days post-vaccination (aHR 0·44; 95% CI 0·24, 0·81) (Table 3). At 35-48 days, the adjusted hazard ratio was similar at 0·38 (95% CI 0·19, 0·77). At 49 or more days, the estimates were much less precise and no longer significantly different to the unvaccinated group (aHR 0·49, 95% CI 0·20, 1·17). In this adjusted model, prior infection was strongly associated with a reduced hazard of subsequent infection (aHR 0·19, 95%CI 0·12, 0·30).

Ct values were available for 1,070 (80·1%) of PCR-positive tests, from 13 laboratories using 6 different validated assays (Table S1). The mean Ct value of 552 PCR-positive tests from the unvaccinated group was 26·6. While no difference was seen when compared with the 411 PCR-positives from between 0-27 days post-vaccination (mean Ct 26·9, p=0·158), the mean Ct value of the 107 PCR-positives occurring 28+ days post-vaccination was significantly higher (mean Ct 31·3, p<0·001) (Figure 2; Table S2). A sensitivity analysis limited to a single assay with the most available results gave the same finding (Table S3).

**Figure 1.**
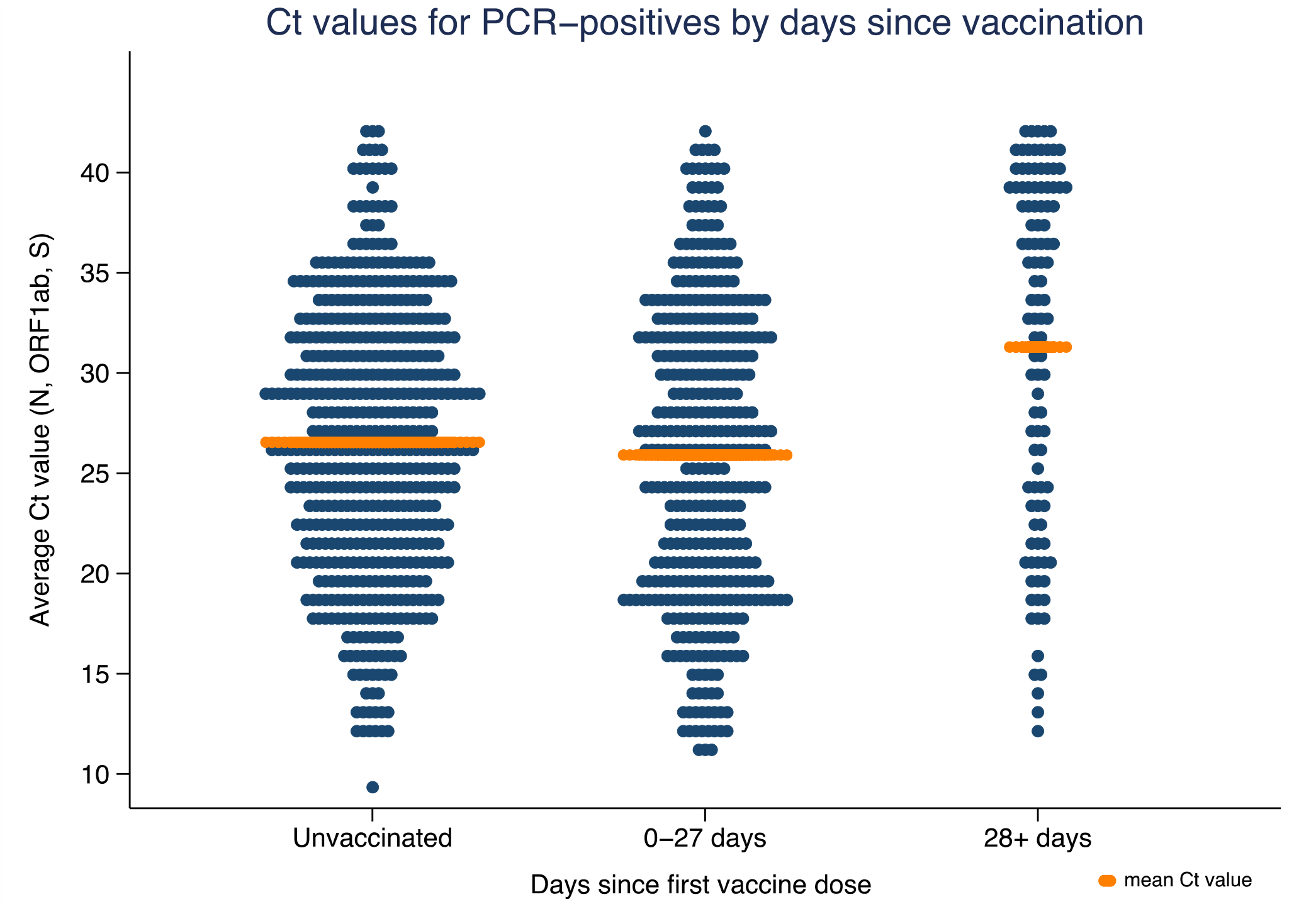
Cycle threshold (Ct) values for PCR-positive infections* by days since vaccination, and the mean cycle threshold value for each category**. *Mean of N, ORF1ab, and S gene target results, according to availability. **Unvaccinated: mean Ct value 26·6; 0-27 days post-vaccination: mean Ct value 26·9 (p=0·158); 28+ days post-vaccination: mean Ct value 31·3 (p<0·001).

**Figure 2.**
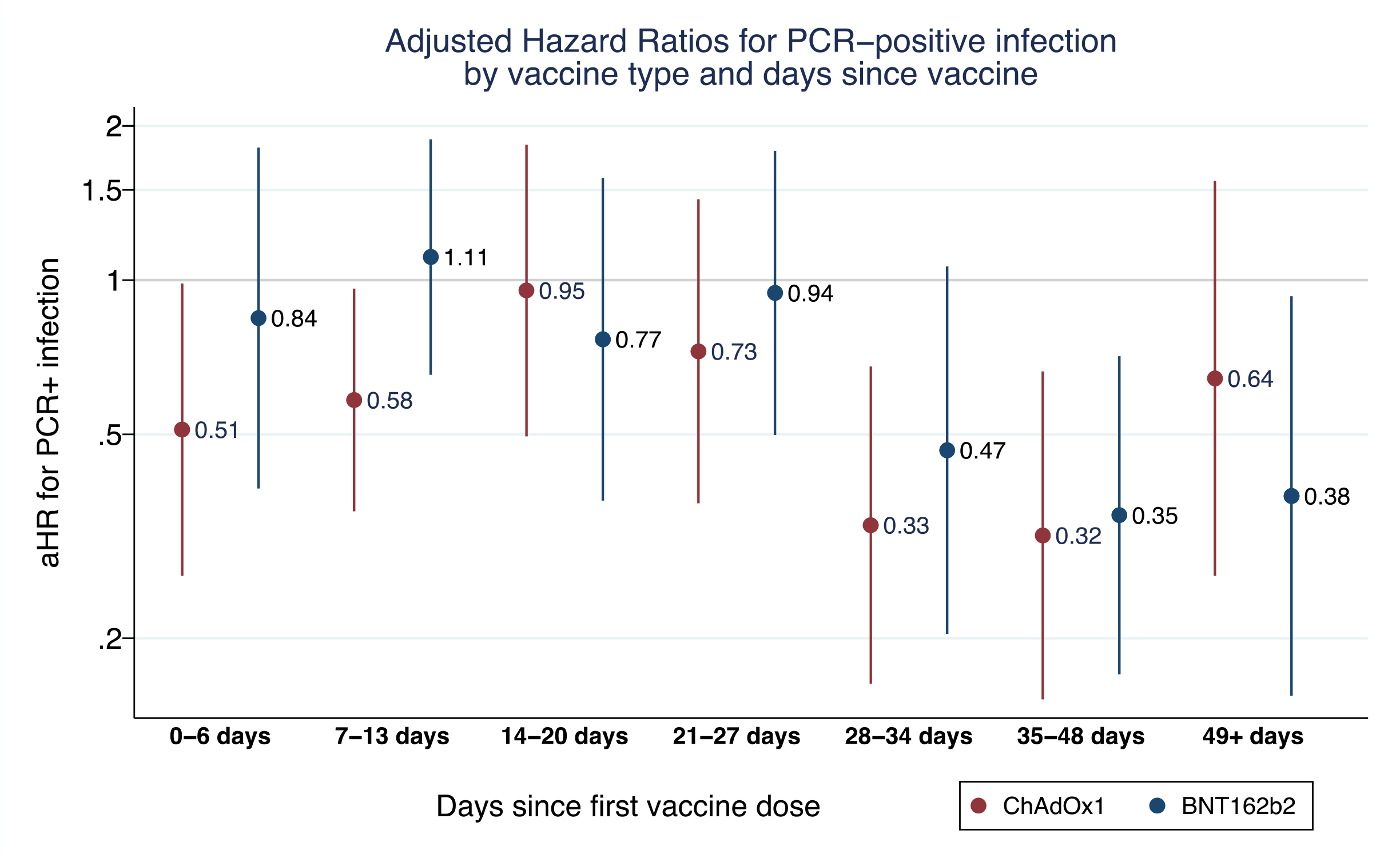
Adjusted hazard ratios* for PCR-positive infection by vaccine type and days since vaccination. * Adjusted hazard ratios for infection estimated using Cox proportional regression model and presented with 95% confidence intervals. Hazard ratios are adjusted for age, sex, prior infection (positive PCR or antibody result) LTCF bed capacity, and local infection incidence rates. 95% confidence intervals are calculated using

### Secondary analyses

The adjusted hazard ratios for infection after ChAdOx1 vaccination were 0·33 (95%CI 0·16, 0·68) at 28-34 days, and 0·32 (95%CI 0·15, 0·66) at 35-48 days post-vaccination; while after BNT162b2 vaccination they were 0·47 (95%CI 0·20, 1·06) at 28-34 days, and 0·35 (95%CI 0·17, 0·71) at 35-48 days post-vaccination (Table 4; Figure 2). A reduced risk of PCR-positive infection was seen in the early post-vaccination period (0-13 days) in residents who received ChAdOx1 (0-6 days: aHR 0·51, 95% CI 0·26-0·99; 7-13 days: aHR 0·58, 95%CI 0·35, 0·96) but not BNT162b.

**Table 4.** Infection rates and adjusted hazards ratios (aHR)* for PCR-confirmed infection compared with unvaccinated group, for the first dose of ChAdOx1 or BNT162b2 vaccine, by days since vaccination.

|  | Person days at risk | Infection events | Infection rate per 10,000 person days | aHR | aHR 95% CI |  | p-value | Person days at risk | Infection events | Infection rate per 10,000 person days | aHR | aHR 95% CI |  | p-value |
| --- | --- | --- | --- | --- | --- | --- | --- | --- | --- | --- | --- | --- | --- | --- |
| Unvaccinated | 338,003 | 723 | 21·39 | 1 | NA | NA | NA | NA | NA | NA | 1 | NA | NA | NA |
| *Vaccine type* | **ChAdOx1** |  |  |  |  |  |  |  | **BNT162b2** |  |  |  |  |  |
| 0-6 days | 30,823 | 61 | 19·79 | 0·51 | 0·26 | 0·99 | 0·045 | 16,768 | 44 | 26·24 | 0·84 | 0·39 | 1·81 | 0·663 |
| 7-13 days | 34,598 | 66 | 19·08 | 0·58 | 0·35 | 0·96 | 0·035 | 18,913 | 73 | 38·60 | 1·11 | 0·65 | 1·88 | 0·700 |
| 14-20 days | 32,672 | 82 | 25·10 | 0·95 | 0·50 | 1·84 | 0·889 | 17,690 | 50 | 28·26 | 0·77 | 0·37 | 1·58 | 0·473 |
| 21-27 days | 30,640 | 50 | 16·32 | 0·73 | 0·37 | 1·44 | 0·358 | 16,874 | 45 | 26·67 | 0·94 | 0·50 | 1·79 | 0·859 |
| 28-34 days | 27,041 | 23 | 8·51 | 0·33 | 0·16 | 0·68 | 0·002 | 16,095 | 19 | 11·80 | 0·47 | 0·20 | 1·06 | 0·070 |
| 35-48 days | 34,705 | 36 | 10·37 | 0·32 | 0·15 | 0·66 | 0·002 | 28,307 | 23 | 8·13 | 0·35 | 0·17 | 0·71 | 0·004 |
| 49+ days | 7,421 | 16 | 21·56 | 0·64 | 0·26 | 1·56 | 0·329 | 20,078 | 24 | 11·95 | 0·38 | 0·15 | 0·93 | 0·034 |
| *Total (vaccine type)* | 197,900 | 334 | 16·88 | NA | NA | NA | NA | 134,725 | 278 | 20·63 | NA | NA | NA | NA |

NA – not applicable
\*Adjusted hazard ratios were estimated using Cox proportional hazards regression according to days since the first vaccine dose for each vaccine type (ChAdOx1 and BNT162b2); the comparator was the unvaccinated group. Hazard ratios are adjusted for age, sex, local monthly infection incidence, and LTCF bed capacity; 95% confidence intervals were calculated using robust standard errors for LTCF-level effects.

The risk of PCR-positive infection was substantially lower in unvaccinated residents with prior infection compared to unvaccinated residents who had not been previously infected (aHR 0·12, 95% CI 0·04-0·35) (Table S4). When compared against unvaccinated residents with no prior infection, unvaccinated residents with prior infection had a significantly lower hazard of infection (aHR 0·12, 95%CI 0·04, 0·35). Amongst those with prior infection, there was no evidence to suggest that a single dose vaccination further reduced the risk of PCR-positive infections at any time point (Table S5).

### Sensitivity analysis

Our sensitivity analysis excluded residents who were never vaccinated despite vaccination occurring in their LTCF, to remove any potential bias due to differences in likelihood of vaccination and risk of exposure, and to explore the effect of herd immunity. 439 (12·0%) of 1,252 never-vaccinated residents had at least one PCR result available >30 days following the date of first vaccination in their LTCF (Table S6), indicating that they remained unvaccinated while other residents were being vaccinated. Removing this group from the analysis reduced the adjusted hazard ratios for infection to 0·29 (95%CI 0·13, 0·63) at 28-34 days following the first dose of vaccination, compared to the unvaccinated group (Table S7).

## Discussion

In this large cohort study of more than 10,400 LTCF residents from across England, single dose vaccination with ChAdOx1 or BNT162b2 was associated with a substantially reduced risk of PCR-positive SARS-CoV-2 infection from 28 days, and this effect was maintained for at least 7 weeks. We estimated vaccine effectiveness to be 56% (19-76%) at 28-34 days, and 62% (23-81%) at 35-48 days. We have only evaluated the effect of the first dose of each vaccine, but our findings constitute the first evidence on the real-world effectiveness of the ChAdOx1 vaccine. Additionally, whereas most trials and observational studies have addressed vaccine effectiveness against symptomatic disease, we show that vaccination reduces the total number of infections (asymptomatic and symptomatic) in frail older adults, and thus transmissibility. An effect of vaccination on transmissibility is further supported by the finding of higher Ct values, implying lower potential for transmission from residents with post-vaccination breakthrough infections compared to unvaccinated residents.

Older adults with comorbidities and frailty have largely been excluded from SARS-CoV-2 vaccine trials, despite the potential for attenuated vaccine responses due to age-related changes in adaptive immune function. While the evidence on immunogenicity of BNT162b2 in older adults is mixed,^24–26^ pooled data from four ChAdOx1 trials, including over 950 participants aged ≥70 years, indicated VE of 63·9% (46-75·9%) against all infection at 22-90 days following a single dose,^17^ which is in line with our findings. The only available data in LTCF residents, from a Danish observational study, suggests that a single dose of BNT162b2 is ineffective in preventing infection,^27^ however participants received the second vaccine dose 24 days following the first dose on average, which, based on our findings, is likely too short to capture the protective effects of a single vaccine dose. Two other observational studies report VE from adults aged over 70-80 years, and report similar or slightly higher estimates from 28-34 days,^15,16^ although results are not directly comparable to our findings as these studies examine VE against symptomatic infection and hospitalisation only, due to the lack of routine testing outside health and care settings.

We observed reduced hazards of infection in the immediate post-vaccination period (0-13 days) for ChAdOx1, which cannot be attributed to protective effects of the vaccine but may be because recently vaccinated LTCFs were less likely to have ongoing outbreaks of infection. Guidance on risk assessment-based deferral of vaccination in LTCFs with active outbreaks was introduced at the end of December 2020,^28^ and this is likely to have disproportionately affected LTCFs predominantly using ChAdOx1, which was deployed later. A similar effect has been observed following single dose vaccination in healthcare workers,^29^ which was attributed to vaccine deferral due to COVID-19 illness. The impact of vaccination deferral on our estimates of VE could not be ascertained because deferral decisions were not routinely recorded and are likely to have varied between settings.

We identified 439 individuals in our cohort who remained unvaccinated despite vaccine roll-out within their LTCF. It is likely that at least a subset of these individuals were receiving end-of-life care, but this cannot be confirmed without accessing primary care records, which were not available for this study. These residents had substantially lower rates of PCR-positive infection than the wider never vaccinated group; consequently, sensitivity analysis excluding this group increased the estimates of vaccine effectiveness to 76% (aHR 0·24, 95%CI 0·09, 0·63) at 35-48 days post-vaccination. This may be attributable to lower risk of exposure in this group, or it may be the result of a broader herd immunity effect following vaccination in the LTCF.

### Strengths and Limitations

A major strength of our analysis is that we were able to access high-quality routine data for a large, well-defined cohort of LTCF residents who were tested regularly for SARS-CoV-2 throughout follow-up. This allowed us to estimate the impact of vaccination on all infections, in contrast to trials and most observational studies which have focused on symptomatic infections. The analysis period coincided with the second wave of the pandemic, making it possible to estimate vaccine effectiveness against infections in the context of rapid emergence of the highly transmissible B.1.1.7 variant. Our cohort included a range of LTCF types therefore we expect these findings to be generalisable across LTCF resident populations. Regarding limitations, as vaccines were rolled out rapidly in LTCFs in England and most resident vaccinations were completed over 1-2 days, we did not attempt to quantify indirect vaccine effects attributable to herd immunity. We were also unable to assess the proportion of LTCFs that deferred vaccination due to outbreaks. Though it is likely that we have underestimated prior infection due to low rates of PCR testing in the first wave of the pandemic, given that prior infection appears highly protective against reinfection we would expect this to attenuate our overall vaccine effectiveness estimates. In addition, we considered Ct values, which correlate with the ability to isolate virus,^30^to be indicative of infectivity. Although it is challenging to compare Ct values across different assays, all results targeted the same genes (N, ORF1ab, S), and similar findings were obtained in sensitivity analysis based on a single assay. Future analyses should also consider outcomes such as hospital admission and mortality, although treatment escalation decisions in the context of end-of-life care are likely to influence vaccination and COVID-19 outcomes.

### Conclusions

Single dose vaccination with either ChAdOx1 or BNT162b reduces the risk of SARS-CoV-2 in frail older residents of LTCFs. Our findings suggest that vaccination also impacts on SARS-CoV-2 transmissibility by reducing the total number of infections in residents, as well as their infectivity. The protective effect of a single dose of vaccination is evident from 4 weeks to at least 7 weeks post-vaccination, which provides some evidence to support extension of the dose interval beyond 3 weeks, in line with UK policy. Further work is required to evaluate the effectiveness of the second dose of the vaccine, and the direct and indirect effects of vaccination against SARS-CoV-2 infection and transmission. This knowledge will be critical to informing policy decisions regarding re-vaccination schedules in this vulnerable population, and the short, medium and long-term disease control measures needed to protect LTCFs from future waves of SARS-CoV-2 infections.

## Data Availability

MS, TP, AC, LS and MK had full access to the data in the study.

## Contributors

Study conceptualisation: LS, AC, JLB, MS, AH, MK, SS. Statistical methodology: AC, TP, MS, LS. Formal analysis: MS, MK, TP. Project administration: MK, BA, CF, AIS. Data curation and validation: MK, DD. Literature review: BA, MK, MS. Funding: LS, AH, AC. Writing (original draft): MS, LS, MK. Writing (review and editing): All authors. MS, TP, AC, LS and MK had full access to the data in the study. LS and AC have shared responsibility for the decision to submit for publication.

## Data Sharing

De-identified test results and limited meta-data will be made available for use by researchers in future studies, subject to appropriate research ethical approvals, once the VIVALDI study cohort has been finalised. These datasets will be accessible via the Health Data Research UK Gateway https://www.healthdatagateway.org/.

## Acknowledgements

The authors would like to thank the staff and residents in the LTCFs that participated in this study, and Mark Marshall at NHS England who pseudonymised the electronic health records. This report is independent research funded by the Department of Health and Social Care (COVID-19 surveillance studies). AH is supported by Health Data Research UK (HDR-UK; grant no LOND1), which is funded by the UK Medical Research Council, Engineering and Physical Sciences Research Council, Economic and Social Research Council, Department of Health and Social Care (England), Chief Scientist Office of the Scottish Government Health and Social Care Directorates, Health and Social Care Research and Development Division (Welsh Government), Public Health Agency (Northern Ireland), British Heart Foundation, and Wellcome Trust. MK is funded by a Wellcome Trust Clinical PhD Fellowship (222907/Z/21/Z). LS is funded by a National Institute for Health Research (NIHR) Clinician Scientist Award (CS-2016-007). The views expressed in this publication are those of the authors and not necessarily those of the NHS, Public Health England, or the Department of Health and Social Care.

## References

1 Eurostat. Beds in residential long-term care facilities. 2020. Available from: https://ec.europa.eu/eurostat/cache/metadata/Annexes/hlth_res_esms_an9.pdf. (Accessed 25 March 2021)

2 European Centers for Disease Control. LTCF data. 2020. Available from: www.ecdc.europa.eu. https://www.ecdc.europa.eu/en/all-topics-z/coronavirus/threats-and-outbreaks/covid-19/prevention-and-control/LTCF-data. (Accessed 25 March 2021).

3 Webster P. COVID-19 highlights Canada’s care home crisis. Lancet (London, England) 2021; 397: 183.

4 The New York Times. More Than One-Third of U.S. Coronavirus Deaths Are Linked to Nursing Homes. 2021. Available from: https://www.nytimes.com/interactive/2020/us/coronavirus-nursing-homes.html. (Accessed 25 March 2021).

5 Department of Health and Social Care. Independent Report - Joint Committee on Vaccination and Immunisation: Advice on priority groups for COVID-19 vaccination, 30 December 2020. Available from: https://www.gov.uk/government/publications/priority-groups-for-coronavirus-covid-19-vaccination-advice-from-the-jcvi-30-december-2020/joint-committee-on-vaccination-and-immunisation-advice-on-priority-groups-for-covid-19-vaccination-30-december-2020. (Accessed 25 March 2021).

6 Medicines and Healthcare products Regulatory Agency. Regulatory approval of COVID-19 Vaccine AstraZeneca. 2020. Available from: https://www.gov.uk/government/publications/regulatory-approval-of-covid-19-vaccine-astrazeneca. (Accessed 25 March 2021).

7 Medicines and Healthcare products Regulatory Agency. Regulatory approval of Pfizer/BioNTech vaccine for COVID-19. 2020. Available from: https://www.gov.uk/government/publications/regulatory-approval-of-pfizer-biontech-vaccine-for-covid-19. (Accessed 25 March 2021).

8 Polack FP, Thomas SJ, Kitchin N, et al. Safety and Efficacy of the BNT162b2 mRNA Covid-19 Vaccine. N Engl J Med 2020; 383: 2603–15.

9 Voysey M, Clemens SAC, Madhi SA, et al. Safety and efficacy of the ChAdOx1 nCoV-19 vaccine (AZD1222) against SARS-CoV-2: an interim analysis of four randomised controlled trials in Brazil, South Africa, and the UK. Lancet 2021; 397: 99–111.

10 Prendki V, Tau N, Avni T, et al. A systematic review assessing the under-representation of elderly adults in COVID-19 trials. BMC Geriatr 2020; 20: 538.

11 Koff WC, Williams MA. Covid-19 and Immunity in Aging Populations — A New Research Agenda. N Engl J Med 2020; 383: 804–5.

12 Gustafson CE, Kim C, Weyand CM, Goronzy JJ. Influence of immune aging on vaccine responses. J Allergy Clin Immunol 2020; 145: 1309–21.

13 Ciabattini A, Nardini C, Santoro F, Garagnani P, Franceschi C, Medaglini D. Vaccination in the elderly: The challenge of immune changes with aging. Semin Immunol 2018; 40: 83–94.

14 Fulop T, Pawelec G, Castle S, Loeb M. Immunosenescence and Vaccination in Nursing Home Residents. Clin Infect Dis 2009; 48: 443–8.

15 Bernal JL, Andrews N, Gower C, et al. Early effectiveness of COVID-19 vaccination with BNT162b2 mRNA vaccine and ChAdOx1 adenovirus vector vaccine on symptomatic disease, hospitalisations and mortality in older adults in the UK: a test negative case control study. MedRxiv 2021. DOI:10.1101/2021.03.01.21252652.

16 Vasileiou E, Simpson CR, Robertson C, et al. Effectiveness of First Dose of COVID-19 Vaccines Against Hospital Admissions in Scotland: National Prospective Cohort Study of 5.4 Million People. SSRN Electron J 2021. DOI:10.2139/ssrn.3789264.

17 Voysey M. Single dose administration and the influence of the timing of booster dose on immunogenicity and efficacy of ChAdOx1 nCoV-19 (AZD1222) vaccine. 2011; 7914.

18 Department of Health and Social Care. Independent report - Optimising the COVID-19 vaccination programme for maximum short-term impact. 2021. Available from: https://www.gov.uk/government/publications/prioritising-the-first-covid-19-vaccine-dose-jcvi-statement/optimising-the-covid-19-vaccination-programme-for-maximum-short-term-impact. (Accessed 25 March 2021).

19 Public Health England. Investigation of novel SARS-CoV-2 variants of concern. 2020. Available from: https://www.gov.uk/government/publications/investigation-of-novel-sars-cov-2-variant-variant-of-concern-20201201. (Accessed 25 March 2021).

20 Krutikov M, Hayward A, Shallcross L. Spread of a Variant SARS-CoV-2 in Long-Term Care Facilities in England. N Engl J Med 2021; : NEJMc2035906.

21 Krutikov M, Palmer T, Donaldson A, et al. Study Protocol: Understanding SARS-Cov-2 infection, immunity and its duration in care home residents and staff in England (VIVALDI). Wellcome Open Res 2021; 5: 232.

22 Department of Health and Social Care. Guidance - Coronavirus (COVID-19) testing in adult care homes. 2021. Available from: https://www.gov.uk/government/publications/coronavirus-covid-19-testing-in-adult-care-homes. (Accessed 25 March 2021).

23 Public Health England. Guidance - COVID-19: management of staff and exposed patients or residents in health and social care setting. 2021. Available from: https://www.gov.uk/government/publications/covid-19-management-of-exposed-healthcare-workers-and-patients-in-hospital-settings/covid-19-management-of-exposed-healthcare-workers-and-patients-in-hospital-settings. (Accessed 25 March 2021).

24 Müller L, Andrée M, Moskorz W, et al. Age-dependent immune response to the Biontech / Pfizer BNT162b2 COVID-19 vaccination. 2021; : 0–14.

25 Collier DA, Ferreira IATM, Datir R, et al. Age-Related Heterogeneity in Neutralising Antibody Responses to SARS-CoV-2 Following BNT162b2 Vaccination. SSRN Electron J 2021. DOI:10.2139/ssrn.3782450.

26 Subbarao S, Warrener LA, Hoschler K, et al. Robust antibody responses in 70 – 80-year-olds 3 weeks after the first or second doses of Pfizer / BioNTech COVID-19 vaccine, United Kingdom, January to February 2021. Eurosurveillance 2021; 26: 1–6.

27 Moustsen-Helms IR, Emborg H-D, Nielsen J, et al. Vaccine effectiveness after 1st and 2nd dose of the BNT162b2 mRNA Covid-19 Vaccine in long-term care facility residents and healthcare workers – a Danish cohort study. MedRxiv 2021.

28 National Health Service. Guidance for COVID-19 vaccination in care homes that have cases and outbreaks. 2020. Available from: https://www.england.nhs.uk/coronavirus/publication/guidance-for-covid-19-vaccination-in-care-homes-that-have-cases-and-outbreaks/. (Accessed 25 March 2021).

29 Hall VJ, Foulkes S, Saei A, et al. Effectiveness of BNT162b2 mRNA Vaccine Against Infection and COVID-19 Vaccine Coverage in Healthcare Workers in England, Multicentre Prospective Cohort Study (the SIREN Study). SSRN Electron J 2021. DOI:10.2139/ssrn.3790399.

30 Singanayagam A, Patel M, Charlett A, et al. Duration of infectiousness and correlation with RT-PCR cycle threshold values in cases of COVID-19, England, January to May 2020. Eurosurveillance 2020; 25. DOI:10.2807/1560-7917.ES.2020.25.32.2001483.

