## Supplementary material for "Vaccine effectiveness of the first dose of ChAdOx1 nCoV-19 and BNT162b2 against SARS-CoV-2 infection in residents of Long-Term Care Facilities (VIVALDI study)"

### Supplementary Materials

I. Additional Methods

**Table S1** – Laboratories and assays for PCR-positive samples with Ct values available

**Figure S1** - Study inclusion flow diagram

II. Cycle threshold values analysis

**Table S2** - Comparison of available mean Ct values by vaccination status

**Table S3** - Comparison of mean Ct values from a single assay (PerkinElmer)

III. Secondary analysis - vaccine effect against infection by prior infection status

**Table S4** - Infection rates and adjusted hazard ratios for those without prior infection

**Table S5** - Infection rates and adjusted hazard ratios for those with prior infection

IV. Sensitivity analysis - vaccine effect against infection excluding never vaccinated sub-group

**Table S6** - Characteristics of never vaccinated residents

**Table S7** - Infection rates and adjusted hazard ratios from sensitivity analysis

V. STROBE Statement

### I. Additional Methods

The COVID-19 national testing programme was rapidly set up in response to the pandemic in April 2020 and consists of five testing Pillars [https://www.gov.uk/government/publications/coronavirus-covid-19-scaling-up-testing-programmes]. Samples from the VIVALDI study are processed under Pillar 1 or Pillar 2 of this programme. Testing for Pillar 1 is undertaken in hospital settings or as part of public health outbreak investigations and samples are processed in NHS hospitals. Pillar 2 encompasses surveillance testing in community settings such as LTCFs and schools. These samples are processed in a national network of accredited laboratories that use a range of PCR-based assays targeting up to three SARS-CoV-2 genes, including N, ORF, and S. The majority of PCR test results included in this analysis were from routine testing, therefore were processed through Pillar 2, and it was only possible to obtain Ct values for Pillar 2 samples.

Table S1. Details of laboratories, assay manufacturers, and gene targets for samples included in Ct values analysis (Pillar 2 only)

| **Laboratory** | **Assay Manufacturer** | **Gene target results available*** | **Gene target results used*** |
|---|---|---|---|
| Newport<br>Charnwood<br>Immensa | PerkinElmer | N, ORF1ab | N, ORF1ab |
| Milton Keynes<br>Glasgow<br>Alderley Park<br>Oncologia<br>University of Birmingham | Thermo Fisher | N, ORF1ab, S | N, ORF1ab, S |
| Health Services Laboratories | | N | N |
| Cambridge | Primer design | ORF1ab | ORF1ab |
| Randox | Randox | ORF1ab, E | ORF1ab |
| Accora Lab Zotz | BGI | ORF1ab | ORF1ab |
| iDNA | ID Solutions | N1, N2 | N1, N2 |

*N/N1/N2 = nucleocapsid protein; ORF1ab = open reading frame 1ab polyprotein; S = spike protein; E = envelope protein

Serum samples were analysed to detect SARS-CoV-2 nucleocapsid IgG using the Abbott ARCHITECT system (Abbott, Maidenhead, UK). An index value of 1·4 is used to define a positive result, in line with manufacturer recommendations. Antibody test results were submitted to NHS England and matched to the NHS number using an algorithm based on participant forename and surname, date of birth, sex and postcode to generate the common pseudo-identifier enabling linkage to PCR test results in the COVID-19 Datastore.

The legal basis for accessing personal data from staff and residents without consent under the General Data Protection Regulations (GDPR) was provided by the Coronavirus (COVID-19): notice under 3(4) of the Health Service (Control of Patient Information) Regulations 2002, which was issued by the UK Government to support the national response to COVID-19 [https://www.gov.uk/government/publications/coronavirus-covid-19-notification-of-data-controllers-to-share-information/coronavirus-covid-19-notice-under-regulation-34-of-the-health-service-control-of-patient-information-regulations-2002-general—2].

Figure S1. Study inclusion flow chart

| Population | | Excluded |
|---|---|---|
| Population records: **n= 41,662** | > | n=1,354 duplicate records |
| Unique individuals: **n= 40,996** | > | n=5,876 with no PCRs recorded |
| Individuals with 1+ PCRs: **n= 35,120** | > | n=5,621 with only 1 PCR |
| Individuals with >1 PCRs: **n= 29,499** | > | n=4,928 with no PCR in analysis period |
| Individuals with PCRs in analysis period: **n= 24,571** | > | n=73 who are not at risk within the analysis period |
| Individuals at risk in analysis period: **n= 24,498** | > | n=13,717 who are staff |
| Residents at risk in analysis period: **n= 10,781** | > | n=360 who are aged <65 years |
| Eligible residents: **n= 10,421** | > | n=9 from LTCF (n=1) with lack of vaccine linkage |
| Final analysis cohort: **n= 10,412** | | |

II. Cycle threshold values

**Table S2**. Mean Ct values from all available PCR-positive results, by days since the first vaccine dose.

| Vaccination status | Samples | Mean Ct value | 95% CI | | p-value |
|---|---|---|---|---|---|
| Unvaccinated | 552 | 26·55 | 26·00 | 27·10 | NA |
| 0-27 days | 411 | 26·91 | 25·19 | 26·62 | 0·158 |
| 28+ days | 107 | 31·29 | 29·62 | 32·96 | <0·001 |

NA – not applicable

**Table S3**. Sensitivity analysis using mean Ct values of results from a single assay* only, by days since the first vaccine dose.

| Vaccination status | Samples | Mean Ct value | 95% CI | | p-value |
|---|---|---|---|---|---|
| Unvaccinated | 246 | 28·70 | 27·96 | 29·45 | NA |
| 0-27 days | 146 | 28·84 | 27·66 | 30·03 | 0·837 |
| 28+ days | 71 | 34·75 | 33·01 | 36·50 | <0·001 |

NA – not applicable

*The single assay for which the greatest total number of results were available was the PerkinElmer SARS-CoV-2 Real-time RT-PCR Assay, used by Newport, Charnwood, and Immensa labs (Table S1). Results from other assays were excluded for this sensitivity analysis.

III. Secondary analysis – vaccine effect against infection by prior infection status

**Table S4.** Adjusted hazard ratios* for PCR-confirmed infection by days since vaccination, in the group with no evidence of prior infection**.

| **Days since first vaccine dose** | **No evidence of prior infection** | | | | | | |
|---|---|---|---|---|---|---|---|
| | Person days at risk | Infection events | Infection rate per 10,000 person days | **Adjusted Hazard Ratio** | **aHR 95% CI** | | **p-value** |
| Unvaccinated | 297,832 | 711 | 23·87 | 1 | NA | NA | NA |
| 0-6 days | 41,548 | 104 | 25·03 | 0·64 | 0·39 | 1·07 | 0·090 |
| 7-13 days | 46,684 | 134 | 28·70 | 0·82 | 0·54 | 1·26 | 0·362 |
| 14-20 days | 43,799 | 122 | 27·85 | 0·90 | 0·54 | 1·52 | 0·701 |
| 21-27 days | 41,229 | 94 | 22·80 | 0·93 | 0·54 | 1·61 | 0·791 |
| 28-34 days | 37,260 | 42 | 11·27 | 0·45 | 0·24 | 0·83 | 0·010 |
| 35-48 days | 54,554 | 54 | 9·90 | 0·36 | 0·18 | 0·73 | 0·005 |
| 49+ days | 23,772 | 39 | 16·41 | 0·49 | 0·20 | 1·17 | 0·108 |
| Total | 586,678 | 1,300 | 22·16 | NA | NA | NA | NA |

NA – not applicable

*Adjusted hazard ratios were estimated using a Cox proportional hazards regression model with an interaction term between vaccination status and prior infection status; the comparator is the unvaccinated group with no evidence of prior infection. Hazard ratios are adjusted for age, sex, local monthly infection incidence, LTCF bed capacity, and for estimates from the group with evidence of prior infection. 95% confidence intervals were calculated using robust standard errors for LTCF-level effects.

**Prior infection was defined by a positive PCR result prior to 8 December 2020 or a positive antibody test prior to vaccination was considered evidence of prior infection.

**Table S5**. Adjusted hazard ratios* for PCR-confirmed infection by days since vaccination, in the group with evidence of prior infection**.

| **Days since first vaccine dose** | **Evidence of prior infection** | | | | | | |
|---|---|---|---|---|---|---|---|
| | Person days at risk | Infection events | Infection rate per 10,000 person days | **Adjusted Hazard Ratio** | **aHR 95% CI** | | **p-value** |
| Unvaccinated | 40,171 | 12 | 2·99 | 1 | NA | NA | NA |
| 0-6 days | 6,043 | 1 | 1·65 | 0·33 | 0·03 | 3·22 | 0·339 |
| 7-13 days | 6,827 | 5 | 7·32 | 1·63 | 0·32 | 8·36 | 0·560 |
| 14-20 days | 6,563 | 10 | 15·24 | 3·82 | 0·90 | 16·32 | 0·070 |
| 21-27 days | 6,285 | 1 | 1·59 | 0·51 | 0·05 | 5·02 | 0·561 |
| 28-34 days | 5,876 | 0 | 0 | 0 | NA | NA | NA |
| 35-48 days | 8,458 | 5 | 5·91 | 1·66 | 0·39 | 7·15 | 0·493 |
| 49+ days | 3,727 | 1 | 2·68 | 0·69 | 0·07 | 7·08 | 0·751 |
| Total | 83,950 | 35 | 4·17 | NA | NA | NA | NA |

NA – not applicable

*Adjusted hazard ratios were estimated using a Cox proportional hazards regression model with an interaction term between vaccination status and prior infection status; the comparator is the unvaccinated group with evidence of prior infection. Hazard ratios are adjusted for age, sex, local monthly infection incidence, LTCF bed capacity, and for estimates from the group with evidence of prior infection. 95% confidence intervals were calculated using robust standard errors for LTCF-level effects.

**Prior infection was defined by a positive PCR result prior to 8 December 2020 or a positive antibody test prior to vaccination was considered evidence of prior infection.

IV. Sensitivity Analysis - VE against infection excluding never vaccinated sub-group

**Table S6**. Characteristics of the never-vaccinated residents, and of the sub-group of never vaccinated residents excluded from the denominator for the sensitivity analysis.

| **Never vaccinated residents** | *n* | *%* |
|---|---|---|
| **Total** | 1,252 | 12·0% |
| Age (years) | Median: 86 | IQR: 80-92 |
| Female sex | 810 | 65·0% |
| Prior infection | 84 | 6·7% |
| LTCFs | 271 | 87·4% |
| PCR positives in analysis period | 328 | 26·2% |
| - Routine PCR testing | 251 | 76·5% |
| - Symptomatic at time of routine testing | 20 | 8·0% |
| **Total never vaccinated with 1+ PCR test >30 days after LTCF first vaccination** | 439 | 35·1% |
| Age (years) | 85 | IQR: 79-91 |
| Female sex | 284 | 64·7% |
| Prior infection | 37 | 8·4% |
| LTCFs | 165 | 53·2% |
| PCR positives in analysis period | 23 | 5·2% |
| - Routine PCR testing | 20 | 87·0% |
| - Symptomatic at time of routine testing | 1 | 5·0% |

**Table S7**. Adjusted hazard ratios* for PCR-confirmed infection in days following vaccination, excluding never vaccinated individuals who continued to be PCR tested**.

| **Days since first vaccine dose** | Person days at risk | Infection events | Infection rate per 10,000 person days | **Adjusted Hazard Ratio** | **aHR 95% CI** | | **p-value** |
|---|---|---|---|---|---|---|---|
| Unvaccinated | 308,217 | 700 | 22·7 | 1 | NA | NA | NA |
| 0-6 days | 47,591 | 105 | 22·1 | 0·54 | 0·32 | 0·92 | 0·024 |
| 7-13 days | 53,511 | 139 | 26·0 | 0·67 | 0·41 | 1·09 | 0·105 |
| 14-20 days | 50,362 | 132 | 26·2 | 0·72 | 0·41 | 1·28 | 0·266 |
| 21-27 days | 47,514 | 95 | 20·0 | 0·64 | 0·34 | 1·24 | 0·187 |
| 28-34 days | 43,136 | 42 | 9·7 | 0·29 | 0·13 | 0·63 | 0·002 |
| 35-48 days | 63,012 | 59 | 9·4 | 0·24 | 0·09 | 0·63 | 0·004 |
| 49+ days | 27,499 | 40 | 14·6 | 0·31 | 0·09 | 1·03 | 0·057 |
| Overall | 640,842 | 1,312 | 20·5 | NA | NA | NA | NA |

NA – not applicable

*Adjusted hazard ratios were estimated using Cox proportional hazards regression according to days since the first vaccine dose. Hazard ratios are adjusted for age, sex, local monthly infection incidence, LTCF bed capacity, and for estimates from the group with evidence of prior infection. 95% confidence intervals were calculated using robust standard errors for LTCF-level effects.

**Excluding individuals who remained unvaccinated despite PCR test results from more than 30 days after vaccination commenced at their LTCF.

I. STROBE Statement

| | Item No | Recommendation | Page No |
|---|---|---|---|
| ***Title and abstract*** | | | |
| | *1* | *(a) Indicate the study's design with a commonly used term in the title or the abstract* | *1,2* |
| | | *(b) Provide in the abstract an informative and balanced summary of what was done and what was found* | *2* |
| ***Introduction*** | | | |
| *Background/ rationale* | *2* | *Explain the scientific background and rationale for the investigation being reported* | *4* |
| *Objectives* | *3* | *State specific objectives, including any prespecified hypotheses* | *5* |
| ***Methods*** | | | |
| *Study design* | *4* | *Present key elements of study design early in the paper* | *5,6* |
| *Setting* | *5* | *Describe the setting, locations, and relevant dates, including periods of recruitment, exposure, follow-up, and data collection* | *5,6* |
| *Participants* | *6* | *a) Cohort study? Give the eligibility criteria, and the sources and methods of selection of participants. Describe methods of follow-up* | *5-7* |
| | | *(b) Cohort study? For matched studies, give matching criteria and number of exposed and unexposed* | *NA* |
| *Variables* | *7* | *Clearly define all outcomes, exposures, predictors, potential confounders, and effect modifiers. Give diagnostic criteria, if applicable* | *5-7* |
| *Data sources/ measurement* | *8** | *For each variable of interest, give sources of data and details of methods of assessment (measurement). Describe comparability of assessment methods if there is more than one group* | *5-7* |
| *Bias* | *9* | *Describe any efforts to address potential sources of bias* | *7* |
| *Study size* | *10* | *Explain how the study size was arrived at* | *7* |

| | | | |
|---|---|---|---|
| *Quantitative variables* | *11* | *Explain how quantitative variables were handled in the analyses. If applicable, describe which groupings were chosen and why* | *7* |
| *Statistical methods* | *12* | *(a) Describe all statistical methods, including those used to control for confounding* | *7* |
| | | *(b) Describe any methods used to examine subgroups and interactions* | *7* |
| | | *(c) Explain how missing data were addressed* | *5-7* |
| | | *(d) Cohort study. If applicable, explain how loss to follow-up was addressed* | *6,7* |
| | | *(e) Describe any sensitivity analyses* | *6* |
| ***Results*** | | | |
| *Participants* | *13** | *(a) Report numbers of individuals at each stage of study? eg numbers potentially eligible, examined for eligibility, confirmed eligible, included in the study, completing follow-up, and analysed* | *7, Fig. S1* |
| | | *(b) Give reasons for non-participation at each stage* | *Fig. S1* |
| | | *(c) Consider use of a flow diagram* | *Fig. S1* |
| *Descriptive data* | *14** | *(a)Give characteristics of study participants (eg demographic, clinical, social) and information on exposures and potential confounders* | *7, Tables 1,2* |
| | | *(b) Indicate number of participants with missing data for each variable of interest* | *7, Table 1, Fig. S1* |
| | | *(c) Cohort study? Summarise follow-up time (eg average and total amount)* | *8, Table 3* |
| *Outcome data* | *15** | *Cohort study? Report numbers of outcome events or summary measures over time* | *8, Table 3* |
| *Main results* | *16* | *(a) Report the numbers of individuals at each stage of the study?eg numbers potentially eligible, examined for eligibility, confirmed eligible, included in the study, completing follow-up, and analysed* | *7, Fig. S1* |
| | | *(b) Give reasons for non-participation at each stage* | *Fig. S1* |

| | | | |
|---|---|---|---|
| | | *(c) Consider use of a flow diagram* | *Fig. S1* |
| *Other analyses* | *17* | *Report other analyses done? eg analyses of subgroups and interactions, and sensitivity analyses* | *8,9, Tables 3,4, Tables S2-7* |
| ***Discussion*** | | | |
| *Key results* | *18* | *Summarise key results with reference to study objectives* | *9* |
| *Limitations* | *19* | *Discuss limitations of the study, taking into account sources of potential bias or imprecision. Discuss both direction and magnitude of any potential bias.* | *10* |
| *Interpretation* | *20* | *Give a cautious overall interpretation of results considering objectives, limitations, multiplicity of analyses, results from similar studies, and other relevant evidence* | *3,9,11* |
| *Generalisability* | *21* | *Discuss the generalisability (external validity) of the study results* | *9,10* |
| ***Other information*** | | | |
| *Funding* | *22* | *Give the source of funding and the role of the funders for the present study and, if applicable, for the original study on which the present article is based* | *2,7* |
